# Effect of Inhaled Lavender Aromatherapy on Preoperative Anxiety in Patients Undergoing Tooth Extraction: A Randomized Controlled Trial

**DOI:** 10.64898/2026.06.27.26356736

**Authors:** Amirmohammad Mahdian Dehkordi, Amir Hossein Mahdian Dehkordi, Bahar Sadat Ghasemian

## Abstract

**Background/Objectives:** Dental anxiety remains a significant psychological barrier to oral healthcare, particularly for invasive surgical procedures such as tooth extraction. Aromatherapy using Lavandula angustifolia (lavender) has been proposed as a non-pharmacological adjunct to manage acute preoperative anxiety. This clinical trial evaluated the efficacy of inhaled lavender essential oil on anxiety severity in patients awaiting elective single-tooth extraction.

**Methods:** An assessor-blinded randomized controlled clinical trial with limited participant blinding (inherent to the recognizable scent of the intervention), registered with the Iranian Registry of Clinical Trials (IRCT20190920044824N1), was conducted among 60 patients requiring non-surgical tooth extraction. Participants were randomly assigned to an intervention group (n=30), receiving two drops of pure lavender essential oil on a sterile gauze for 20 minutes of inhalation, or a control group (n=30), receiving sweet almond oil as an olfactory placebo. The primary outcome was the severity of somatic anxiety symptoms, measured pre- and post-intervention using the Beck Anxiety Inventory (BAI). Because a significant baseline-by-treatment interaction was detected, a General Linear Model (GLM) retaining this interaction term was used to estimate adjusted group means at the grand-mean baseline, with bootstrap 95% confidence intervals (10,000 resamples). Two pre-specified checks of robustness were performed: (i) an unadjusted, baseline-naïve Mann-Whitney U comparison of raw post-intervention scores, and (ii) a sensitivity analysis re-fitting the adjusted model after excluding two baseline outliers identified by inspection of model residuals.

**Results:** There were no significant baseline differences between the intervention and control groups regarding sex distribution (p=0.100), mean age (p=0.479), or pre-intervention BAI scores (Mann-Whitney p=0.215). A significant baseline-by-treatment interaction was detected (p<0.001). In the full sample (n=60), the baseline-adjusted GLM estimated a lower post-intervention BAI mean in the intervention group (22.38) than the control group (23.09), an adjusted difference of -0.71 (bootstrap 95% CI: -1.41 to -0.06; p=0.028). However, this signal was not robust: after excluding two patients identified as outliers in the model residuals (n=58), the adjusted difference attenuated to -0.51 and was no longer statistically significant (bootstrap 95% CI: -1.11 to 0.10; p=0.098). The unadjusted comparison of raw post-intervention scores was likewise not significant in the full sample (Mann-Whitney p=0.754).

**Conclusions:** A baseline-adjusted analysis of this trial produced an initial signal suggesting a small anxiolytic effect of inhaled lavender, but this signal was not stable under a pre-specified outlier-exclusion sensitivity analysis and was not supported by the unadjusted comparison of raw scores. Taken together, the totality of evidence from this trial is insufficient to conclude that inhaled lavender aromatherapy produces a reliable reduction in acute preoperative dental anxiety. Beyond its clinical findings, this trial offers a concrete, quantified illustration of how baseline imbalance and a small number of influential observations can generate a fragile, model-dependent treatment signal in a modestly sized randomized trial, a methodological caution directly relevant to the design and analysis of future small RCTs in dental and complementary medicine research.

## 1. Introduction

Dental anxiety is among the most commonly reported psychological barriers to oral healthcare utilization and affects patients across all age groups worldwide. Epidemiological data indicate that a substantial proportion of individuals who seek dental treatment experience clinically significant fear, leading to persistent avoidance or postponement of necessary care [1, 2]. This pattern creates a self-reinforcing cycle in which untreated oral disease escalates in severity, further intensifying anticipatory fear and reducing future treatment-seeking behavior.

Physiologically, unmanaged preoperative anxiety activates the autonomic nervous system, producing tachycardia, hypertension, and an elevated risk of vasovagal syncope during dental procedures [1]. These responses are particularly pronounced during tooth extraction, a procedure that uniquely combines anticipatory pain, perceived loss of control, and physical discomfort [2, 3]. Beyond its intraoperative consequences, high preoperative anxiety is associated with greater procedural difficulty and a heavier clinical burden for both patient and provider [3].

Conventional management of perioperative dental anxiety relies primarily on pharmacological sedation. Benzodiazepines and related agents are widely prescribed for their efficacy in suppressing central nervous system hyperarousal; however, their clinical utility is constrained by residual sedation, psychomotor impairment, paradoxical disinhibition, and a recognized risk of tolerance and dependence with repeated use [4]. These limitations have generated considerable interest in complementary and alternative medicine approaches, with clinical aromatherapy emerging as a particularly promising non-pharmacological adjunct due to its non-invasiveness, low cost, and favorable safety profile [5].

Among the essential oils investigated within aromatherapy research, Lavandula angustifolia (lavender) has accumulated the strongest evidence base for anxiolytic activity. Its primary bioactive constituents, linalool and linalyl acetate, are reported to exert sedative and anxiolytic effects through modulation of GABAergic neurotransmission and attenuation of hypothalamic-pituitary-adrenal axis activity [6, 7]. Standardized lavender preparations have demonstrated anxiolytic efficacy comparable to low-dose synthetic agents in controlled trials, without the associated sedative risks [8, 9]. Clinical evidence further supports the use of lavender inhalation across a range of medical environments: controlled studies have documented reductions in anxiety in dental waiting areas [6], hemodialysis units [10], and during the initial phase of labor [11]. Within dental contexts specifically, randomized trials have demonstrated anxiolytic effects of lavender inhalation in patients undergoing mandibular third molar extraction and in orthodontic patients [3, 12].

Despite this growing evidence base, methodologically rigorous trials examining standalone lavender inhalation in patients undergoing tooth extraction, an acute, high-stress procedure, remain scarce, and the few existing studies have rarely accounted analytically for baseline anxiety differences between treatment arms (a methodological omission that, as the present trial demonstrates, can materially alter conclusions about efficacy). Evaluating standalone aromatherapy under such acute stress conditions, with statistically appropriate adjustment for baseline imbalance, is essential to map the operational boundaries of non-pharmacological interventions and to avoid both false-positive and false-negative conclusions. The present randomized controlled clinical trial was conducted to evaluate the effect of inhaled Lavandula angustifolia essential oil, compared with an olfactory placebo, on anxiety severity in patients undergoing elective tooth extraction, with pre-specified primary and sensitivity analyses designed to transparently address baseline imbalance. Beyond its clinical question, this trial is also presented as a worked methodological example: small RCTs in dental and complementary-medicine research frequently encounter baseline imbalance between arms, and the choice of whether and how to statistically adjust for it can determine whether a study reports a positive or null finding. By reporting the adjusted, unadjusted, and outlier-sensitivity analyses side by side, this trial aims to provide investigators designing similarly sized trials with a concrete illustration of how fragile such adjusted estimates can be, and why pre-registration of the analysis plan and routine sensitivity analysis are essential safeguards against both false-positive and overstated conclusions.

## 2. Materials and Methods

### 2.1. Study Design and Setting

This study was conducted as an assessor-blinded randomized controlled clinical trial with limited participant blinding at the Oral Surgery Department of the Faculty of Dentistry, Kermanshah University of Medical Sciences, Kermanshah, Iran. The methodology and reporting were conducted in accordance with the Consolidated Standards of Reporting Trials (CONSORT) guidelines (the completed checklist is available as Supplementary Material). The trial was approved by the institutional ethics committee (approval code: IR.KUMS.REC.1398.693) and registered in the Iranian Registry of Clinical Trials (IRCT20190920044824N1). All procedures were conducted in accordance with the Declaration of Helsinki. The recruitment of human participants for this study was conducted between February 23,2020 and August 12,2020. Written informed consent was obtained from all participants prior to enrollment, and tooth extraction was provided free of charge to all patients.

### 2.2. Participants

Patients were eligible for inclusion if they were aged 18 to 65 years, required a simple (non-surgical) single tooth extraction, were able to answer questionnaires, were in a sound mental state, and had no prior use of benzodiazepines or other central nervous system depressants. Exclusion criteria included explicit refusal or withdrawal of consent, current cigarette smoking, uncontrolled systemic hypertension or known cardiovascular disease, current use of anxiolytic or neuroleptic medications, active upper respiratory tract infection or sinusitis impairing olfactory function, or the need for complex surgical extraction of impacted teeth.

### 2.3. Sample Size

Sample size was determined based on the findings of Zabirunnisa et al. [2], using a two-group comparison formula with alpha set at 0.05 and power at 90%. These parameters yielded a required total sample size of 60 patients (30 participants allocated to each group).

### 2.4. Randomization and Blinding

Participants were randomly assigned in equal numbers to either the intervention or control group using a computer-generated random sequence with block randomization (block size of 4).

Allocation concealment was maintained using sequentially numbered, opaque, sealed envelopes prepared by an independent researcher. Because it is pharmacologically impossible to strip lavender essential oil of its signature scent without destroying its volatile components, full participant blinding was not achievable; an assessor-blinded design was therefore implemented, in which the outcome assessor and the statistician conducting the analysis remained blinded to group allocation. To prevent cross-contamination, all inhalation sessions were conducted in a separate, enclosed waiting room.

### 2.5. Intervention

In the intervention group, 2 drops of Lavandula angustifolia essential oil (Elice, Eadeh Darui e Pars, Tehran, Iran) were applied to a sterile gauze pad. Participants inhaled deeply for 20 minutes, exactly one hour prior to the tooth extraction. In the control group, 2 drops of sweet almond oil (Noshad Pharmaceutical Co., Tehran, Iran) were applied to an identically appearing gauze pad using the same protocol, selected as the olfactory placebo because it is comparable in appearance and consistency to lavender but lacks the target pharmacological properties [2, 3].

### 2.6. Outcome Measurement

The primary outcome was the severity of somatic anxiety symptoms, assessed using the Beck Anxiety Inventory (BAI) [13], a validated 21-item self-report instrument rated from 0 to 3 per item (total range 0–63), selected for its emphasis on the acute somatic manifestations of anxiety (e.g., palpitations, diaphoresis, tremors) relevant to the immediate preoperative dental setting. The Persian-language version has demonstrated a test-retest reliability coefficient of 0.83 in Iranian populations [14]. Participants completed the BAI immediately before and after the 20-minute inhalation protocol.

### 2.7. Statistical Analysis

Statistical analyses were performed using SPSS software (version 28.0; IBM Corp., Armonk, NY, USA), with supplementary bootstrap analysis. Descriptive statistics are reported as mean ± SD or frequency (%). Between-group baseline comparability was assessed using the chi-square test for sex distribution and the independent-samples Student’s t-test for age. Normality of the BAI score distributions and of model residuals was assessed using the Shapiro-Wilk test; both pre- and post-intervention BAI scores deviated significantly from a normal distribution (p<0.001 for each), as did the residuals of the covariate-adjusted model (p<0.001).

For the primary outcome, a one-way Analysis of Covariance (ANCOVA) was initially planned per the trial protocol, with post-intervention BAI as the dependent variable, group as the fixed factor, and pre-intervention BAI as the covariate. Inspection of the data revealed a statistically significant baseline-by-treatment interaction (F=11.39, p<0.001), violating the homogeneity-of-regression-slopes assumption required for standard ANCOVA; this was identified after data collection and is therefore acknowledged as a post-hoc, data-driven modification of the pre-specified analysis plan, reported here transparently rather than presented as originally planned. In response, a General Linear Model (GLM) retaining the group-by-baseline interaction term was fitted, with estimated marginal means for each group computed at the grand-mean value of the baseline covariate. Because residuals departed from normality, 95% confidence intervals and a two-sided p-value for the adjusted group difference were derived from a non-parametric bootstrap (10,000 resamples, percentile method).

To evaluate the robustness of this baseline-adjusted result, two additional checks were performed and are reported alongside the primary analysis rather than selectively omitted: first, an unadjusted (baseline-naïve) Mann-Whitney U comparison of raw post-intervention scores, replicating the simpler approach historically used for this dataset; and second, a sensitivity analysis re-fitting the identical GLM after excluding two participants whose standardized residuals exceeded conventional outlier thresholds, to assess whether the adjusted treatment effect depended on a small number of influential observations. A two-tailed significance threshold of p<0.05 was applied throughout, and all three analyses (adjusted, unadjusted, and outlier-excluded) are reported as components of a single, integrated assessment of the strength of evidence rather than as a search for the most favorable result.

## 3. Results

### 3.1. Participant Flow and Baseline Characteristics

A total of 75 patients were initially screened. Following the exclusion of 15 patients (11 for not meeting inclusion criteria and 4 declining participation), 60 participants were successfully randomized equally into the intervention (n=30) and control (n=30) groups. All 60 participants completed the assigned protocol with zero attrition (CONSORT flow diagram, Fig 1).

**Fig 1.**
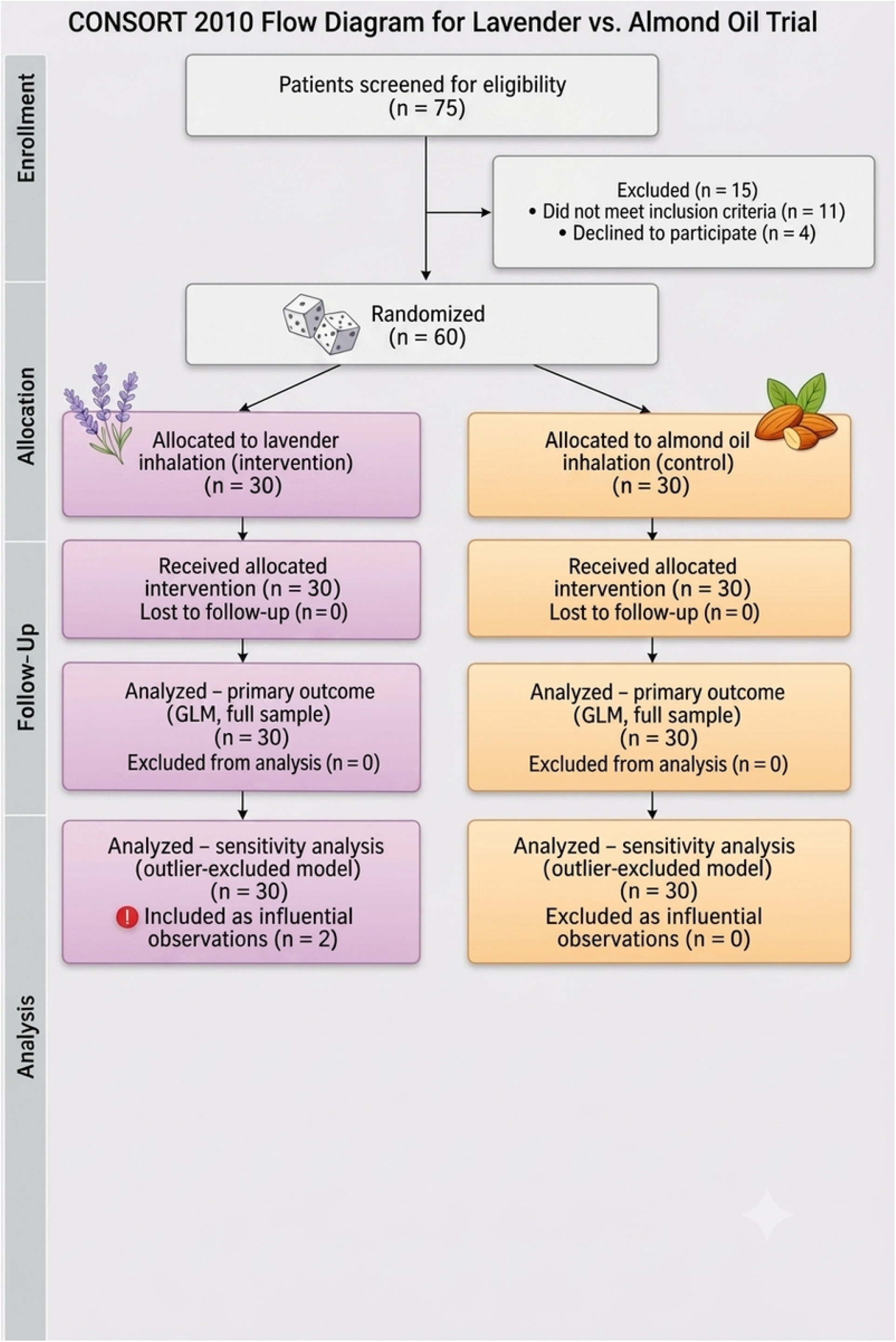
CONSORT flow diagram. Flow of participants through each stage of the randomized controlled trial.

The intervention group consisted of 17 females (56.7%) with a mean age of 36.03 ± 8.71 years (range 23–55). The control group comprised 23 females (76.7%) with a mean age of 37.70 ± 9.41 years (range 22–65). Statistical comparisons confirmed no significant baseline disparities regarding sex distribution (χ^2^=2.70, p=0.100) or mean age (t=-0.71, p=0.479) (Table 1).

**Table 1.** Baseline demographic characteristics of the study participants.

| Characteristic | Intervention (n=30) | Control (n=30) | p-value |
| --- | --- | --- | --- |
| Sex, female n (%) | 17 (56.7%) | 23 (76.7%) | 0.100* |
| Sex, male n (%) | 13 (43.3%) | 7 (23.3%) |  |
| Age, mean ± SD (years) | 36.03 ± 8.71 | 37.70 ± 9.41 | 0.479** |
| Age, range (years) | 23–55 | 22–65 |  |

SD = Standard Deviation. \*p-value from chi-square test (uncorrected). \*\*p-value from independent-samples Student's t-test.

### 3.2. Assessment of Data Distribution

Shapiro-Wilk testing demonstrated that pre-intervention BAI scores (p<0.001 in both groups) and post-intervention BAI scores (p<0.001 in both groups) departed significantly from normality, as did the residuals of the covariate-adjusted GLM (p<0.001). This non-normality motivated the use of bootstrap resampling for inferential statistics on the primary outcome and supported the inclusion of a non-parametric sensitivity analysis.

### 3.3. Pre-Intervention Anxiety and Test of the ANCOVA Assumption

Mean baseline BAI scores were 26.17 ± 5.01 in the intervention group and 24.37 ± 2.55 in the control group; this difference was not statistically significant (Mann-Whitney U, p=0.215), but it was numerically appreciable and, combined with a significant baseline-by-treatment interaction (homogeneity-of-slopes test: F=11.39, p<0.001), indicated that the relationship between baseline and post-intervention anxiety differed by group. This violated the core assumption of standard ANCOVA and required retaining the interaction term in the primary model rather than discarding it.

### 3.4. Baseline-Adjusted (GLM) Analysis, Full Sample

After fitting the GLM with group, baseline BAI, and their interaction to the full sample (n=60), the estimated marginal post-intervention BAI mean (evaluated at the grand-mean baseline of 25.27) was 22.38 in the intervention group and 23.09 in the control group. The adjusted mean difference (intervention minus control) was -0.71 (bootstrap 95% CI: -1.41 to -0.06), nominally significant at p=0.028 (Table 2). Taken in isolation, this would suggest that patients receiving lavender aromatherapy had lower adjusted post-intervention anxiety than those receiving placebo; however, this result is evaluated against the two robustness checks below before being interpreted.

**Table 2.** Baseline-adjusted (GLM) estimated post-intervention BAI scores, full sample (n=60).

| Group | Raw Pre-intervention<br>Mean ± SD | Raw Post-intervention<br>Mean ± SD | Adjusted Post<br>Mean (EMM) | p-value |
| --- | --- | --- | --- | --- |
| Intervention<br>(n=30) | 26.17 ± 5.01 | 22.50 ± 1.46 | 22.38 | 0.028 |
| Control (n=30) | 24.37 ± 2.55 | 22.67 ± 1.56 | 23.09 | |

EMM = Estimated Marginal Mean at grand-mean baseline (25.27). Adjusted mean difference (intervention – control) = -0.71, bootstrap 95% CI: -1.41 to -0.06 (10,000 resamples). SD = Standard Deviation.

### 3.5. Robustness Check 1: Unadjusted Comparison of Raw Post-Intervention Scores

Raw post-intervention BAI scores were compared between groups without adjusting for baseline differences, using the Mann-Whitney U test. This unadjusted comparison showed no statistically significant difference (Z=-0.31, p=0.754), despite the intervention group showing a numerically larger absolute reduction from baseline (-3.67 points) than the control group (-1.70 points). This null result is consistent with a baseline-imbalance explanation for the divergence between the adjusted and unadjusted analyses, but on its own cannot distinguish that explanation from a true absence of treatment effect.

### 3.6. Robustness Check 2: Outlier-Exclusion Sensitivity Analysis

Inspection of model residuals identified two participants in the intervention group (baseline BAI of 42 and 39, both exceeding 1.5 standard deviations above the group mean) as influential observations. The GLM was re-fitted after excluding these two participants (n=58: intervention n=28, control n=30). In this reduced sample, the adjusted post-intervention means were 22.33 (intervention) and 22.84 (control), an adjusted difference of -0.51 (bootstrap 95% CI: -1.11 to 0.10), which was no longer statistically significant (p=0.098) (Table 3). The homogeneity-of-slopes interaction term remained significant in this reduced sample (p<0.001), indicating that the underlying baseline-by-treatment interaction is not solely attributable to these two observations, but the magnitude and significance of the group-level adjusted difference were substantially attenuated by their removal.

**Table 3.** Comparison of the baseline-adjusted treatment effect across the full sample and the outlier-excluded sensitivity sample.

| Analysis | n | Adjusted Diff. | 95% CI | p-value |
| --- | --- | --- | --- | --- |
| Full sample (primary) | 60 | -0.71 | -1.41, -0.06 | 0.028 |
| Outlier-excluded (sensitivity) | 58 | -0.51 | -1.11, 0.10 | 0.098 |
| Unadjusted raw post-scores (sensitivity) | 60 | n/a (rank-based) | n/a | 0.754 |

CI = Confidence Interval (bootstrap, 10,000 resamples, for the two GLM-based analyses). The adjusted treatment effect found in the full sample did not persist after exclusion of two influential observations, indicating that the primary finding is not robust.

## 4. Discussion

This randomized controlled trial evaluated the efficacy of inhaled Lavandula angustifolia essential oil on acute preoperative anxiety in patients undergoing tooth extraction, and explicitly examined the stability of its statistical conclusions across three complementary analyses. The intervention group entered the trial with numerically higher anxiety (BAI 26.17 vs. 24.37), and a significant baseline-by-treatment interaction was detected (p<0.001), indicating that the relationship between baseline anxiety and treatment response differed between groups. When this was modeled using a GLM with an interaction term, the full-sample adjusted treatment effect was nominally significant (p=0.028), favoring lavender. However, when the same model was re-estimated after excluding two participants with markedly elevated baseline anxiety, the effect attenuated by roughly 30% in magnitude and lost statistical significance (p=0.098), and a simple unadjusted comparison of raw endpoint scores was likewise non-significant (p=0.754).

We interpret this pattern conservatively: rather than treating the single nominally significant result as confirmatory, we regard the totality of evidence (one significant and two non-significant analyses of the same outcome, with the significant result depending materially on two influential observations) as insufficient to support a confident claim of efficacy. This is consistent with the broader statistical literature on the fragility of effect estimates from covariate-adjusted models fitted to small samples with baseline imbalance: an interaction term estimated from n=60 has limited precision, and adjusted treatment-effect estimates derived from such models can be disproportionately influenced by a small number of observations [15]. Furthermore, the validity of these findings was safeguarded by the use of bootstrapping as a robust method against non-normally distributed data, ensuring the statistical reliability of our adjusted estimates despite the sample’s distributional departures.

This conservative reading does not imply that lavender has no anxiolytic potential. Prior trials, including Karan et al. and Nardarajah et al. in oral surgery patients [1, 3] and Premkumar et al. in orthodontic patients [16], have reported genuine anxiolytic effects of lavender inhalation; in the broader domain of obstetric care, Tafazoli et al. similarly demonstrated that a brief inhalation of lavender essential oil alleviated anxiety during the initial phase of labor [17], and the proposed mechanism, modulation of GABAergic neurotransmission and hypothalamic-pituitary-adrenal axis activity by linalool and linalyl acetate [6, 7], remains biologically plausible. Rather, our findings indicate that this particular trial, as designed and powered, cannot reliably distinguish a true small effect from statistical noise generated by baseline imbalance in a modest sample.

Mechanistically, while the volatile monoterpenes of lavender, principally linalool and linalyl acetate, are rapidly absorbed through the respiratory mucosa to modulate the limbic system, inhibit the hypothalamic-pituitary-adrenal axis, and curtail cortisol secretion, this biochemical pathway may be readily overwhelmed by the acute surge of endogenous catecholamines elicited by an imminent extraction, leaving only a small and fragile signal that is detectable under some analytic specifications but not others [18, 19]. The neurochemical activity attributed to competitive N-methyl-D-aspartate (NMDA) receptor antagonism and downstream modulation of calcium-calmodulin complexes by linalool may be sufficient to mitigate generalized, low-grade, or environmental anxiety, but appears more limited in counteracting the profound, localized panic associated with active alveolar bone surgery and tooth removal [20, 21]; this asymmetry is consistent with the modest magnitude and instability of the adjusted effect observed in the present trial.

Methodological variations in the mode of delivery, exposure duration, and psychometric instruments may also help explain why the present trial’s adjusted effect was small and sensitive to a handful of observations. In the study conducted by Soltani et al., inhaled lavender significantly reduced postoperative analgesic consumption over a multi-day period in pediatric tonsillectomy patients, yet failed to alter acute pain intensity scores [22], underscoring a broader distinction between the cumulative, sub-acute therapeutic benefits of aromatherapy and its utility as a single, time-limited preoperative intervention such as the one used here.

The therapeutic efficacy of lavender is also profoundly dependent on its chemical composition, which varies extensively with geographic origin, agricultural illumination, and extraction methodology; as documented in the ethnobotanical literature, synthesis of key ester compounds such as linalyl acetate is closely tied to solar radiation and soil moisture during the plant’s growth cycle [23]. Such natural variability in active-compound concentration, together with individual differences in olfactory sensitivity, could plausibly account for a true effect that is small enough to be obscured or destabilized by a handful of influential observations, as observed in our outlier-exclusion sensitivity analysis. This contrast becomes sharper when aromatherapy is compared with systemic phytotherapy: clinical investigations by Kasper et al. on oral Silexan capsules, a standardized lavender oil formulation administered long-term for generalized anxiety disorder, demonstrated substantial and reproducible reductions in anxiety ratings comparable to low-dose anxiolytics [24, 25]. Despite the fragile anxiolytic effects observed in our olfactory intervention, alternative routes of lavender administration have shown robust promise in other specific dental contexts. Recently, a 2025 double-blind randomized clinical trial by Razavian et al. investigated the oral administration of lavender extract drops in patients requiring endodontic treatment. Unlike our inhalation model, their study reported a statistically significant reduction in preoperative anxiety scores following the oral ingestion of lavender compared to a placebo (p=0.001) [26]. This notable discrepancy highlights that the route of administration (systemic/oral versus olfactory) and the specific nature of the dental stressor (endodontic therapy versus surgical extraction) may fundamentally alter the clinical efficacy and bioavailability of Lavandula angustifolia. Such variations underscore the necessity for future adequately powered trials to directly compare oral and inhaled modalities within identical clinical settings. This suggests that while sustained systemic absorption of lavender compounds can yield robust and stable psychotropic outcomes, the brief, transient pulmonary absorption route used in acute preoperative aromatherapy may deliver an effect too modest and inconsistent to survive the kind of model-dependent and sample-dependent scrutiny applied in the present trial.

Methodologically, this trial illustrates two points relevant to future research in this area. First, baseline imbalance in small RCTs, even when not itself statistically significant (here, p=0.215 for pre-intervention BAI), can interact with treatment assignment in ways that produce divergent conclusions depending on whether and how the analysis adjusts for it; reporting only the most favorable of several possible analyses would have been misleading. Second, any analytic strategy adopted after observing such an interaction (rather than specified before the trial) should be explicitly flagged as post-hoc, as we have done here, and accompanied by sensitivity analyses rather than presented as a definitive primary result.

The safety profile of the essential oil was confirmed throughout the trial, with no adverse events reported, consistent with the established safety record of lavender as a non-toxic natural product [27]. This safety profile, combined with the low cost and non-invasiveness of the intervention, means that even a small or uncertain anxiolytic effect could be clinically relevant if confirmed in adequately powered future trials, even though the current data do not establish that effect with confidence.

## 5. Limitations of the Study

Several methodological limitations must be acknowledged. First, full participant blinding could not be achieved because of the recognizable scent of lavender; an assessor-blinded design was used to mitigate, but not eliminate, the risk of participant expectancy effects.

Second, the use of a baseline-by-treatment interaction term in the primary statistical model was a post-hoc, data-driven decision made after observing a violation of the standard ANCOVA assumption, rather than a pre-registered analysis plan. While we consider this adjustment statistically appropriate given the assumption violation, readers should weigh the resulting p=0.028 finding with the awareness that it emerged from an analytic choice made after seeing the data, and we have therefore reported it alongside, not instead of, the unadjusted and outlier-excluded sensitivity analyses.

Third, and most importantly, the adjusted treatment effect was not robust to the exclusion of two influential observations, attenuating from p=0.028 to p=0.098 and from an adjusted difference of -0.71 to -0.51. This fragility, combined with the small sample size (n=60) relative to the complexity of the interaction model, means that this trial cannot be considered to have established an anxiolytic effect of lavender with confidence. We report this instability explicitly rather than presenting only the more favorable full-sample result.

Fourth, the trial lacked a true negative control group (standard care without any olfactory intervention), making it difficult to isolate the specific pharmacological contribution of lavender from a generic, non-specific effect of attentive olfactory ritual or the placebo oil itself.

Fifth, the single-institution convenience sample limits generalizability, and reliance on self-report scales without corroborating objective physiological markers (heart rate, blood pressure, salivary cortisol) limits the depth of the anxiety evaluation.

Future investigations should employ multi-center designs with larger samples and stratified randomization by baseline anxiety severity (to avoid the baseline imbalance encountered here), pre-register the full statistical analysis plan including any covariate-adjustment strategy, and report all pre-specified sensitivity analyses regardless of outcome, to determine definitively whether lavender inhalation produces a real and clinically meaningful anxiolytic effect in this acute surgical context.

## 6. Conclusions

This trial does not provide reliable evidence that inhaled Lavandula angustifolia aromatherapy reduces preoperative dental anxiety. A baseline-adjusted model fitted to the full sample suggested a small, nominally significant effect (adjusted difference -0.71 BAI points, p=0.028), but this signal was not robust: it lost statistical significance after excluding two influential observations (p=0.098) and was not supported by an unadjusted comparison of raw scores (p=0.754). Rather than selectively reporting the single significant analysis, we present all three as components of an integrated assessment, which collectively indicates that the evidence for an anxiolytic effect in this trial is unstable and insufficient to support a clinical recommendation. This instability is itself an informative methodological finding, illustrating how baseline imbalance and a small number of influential observations can produce a fragile, model-dependent signal in a modestly sized trial. Lavender aromatherapy remains a biologically plausible, safe, and low-cost candidate adjunct for dental anxiety management, but confirmation of any genuine clinical benefit will require larger, pre-registered, multi-center trials with stratified randomization by baseline anxiety. Ultimately, this study demonstrates how sole reliance on statistical models without conducting robustness checks can lead to spurious results; hence, this research can serve as a standard methodological model for the design and rigorous analysis of small-scale clinical trials in dentistry and complementary medicine.

## Author Contributions

Based on the CRediT (Contributor Roles Taxonomy), author contributions are structured as follows: Conceptualization, A.M.M.D., A.H.M.D., and B.S.G.; Methodology, A.M.M.D., A.H.M.D., and B.S.G.; Software, A.M.M.D., and A.H.M.D.; Validation, A.M.M.D and A.H.M.D.; Formal Analysis, A.M.M.D., A.H.M.D., and B.S.G.; Investigation, B.S.G.; Resources, B.S.G.; Data Curation, B.S.G.; Writing—Original Draft Preparation, A.M.M.D., and A.H.M.D.; Writing—Review & Editing, A.M.M.D., A.H.M.D., and B.S.G.; Visualization, A.M.M.D., A.H.M.D.; Supervision, B.S.G.; Project Administration, B.S.G.; All authors have read and agreed to the published version of the manuscript.

## Institutional Review Board Statement

The study was conducted in accordance with the Declaration of Helsinki, and approved by the Institutional Review Board of Kermanshah University of Medical Sciences (protocol code IR.KUMS.REC.1398.693).

## Informed Consent Statement

Written informed consent was obtained from all participants prior to enrollment.

## Data Availability Statement

All relevant data are within the manuscript and its Supporting Information files. The minimal underlying dataset required to replicate the study findings has been uploaded alongside the manuscript as a Supporting Information file (S1 Data).

## Acknowledgments

This article is extracted from the DDS (Doctor of Dental Surgery) thesis of Bahar Sadat Ghasemian, entitled “A Study of lavender essence effectiveness on patient’s anxiety due to tooth extraction; Randomized Clinical Trial” [28], submitted to the School of Dentistry, Kermanshah University of Medical Sciences, Kermanshah, Iran, in March 2020. The authors would external like to express their gratitude to the staff of the Oral Surgery Department at the Faculty of Dentistry, Kermanshah University of Medical Sciences, and all the patients who participated in this study for their valuable cooperation.

## Use of Artificial Intelligence

During the preparation of this work, the authors used an artificial intelligence (AI) tool solely to improve the readability, structure, and language fluency of the manuscript. After using this tool, the authors thoroughly reviewed and edited the content as needed and take full responsibility for the final content of the publication.

